# Echocardiographic characterisation in critical Covid19 - an observational study

**DOI:** 10.1101/2021.07.23.21261025

**Authors:** Henrik Isackson, Anders Larsson, Miklos Lipcsey, Robert Frithiof, Frank A. Flachskampf, Michael Hultström

## Abstract

**Objective:** We aimed to investigate the acute cardiac effects of severe SARS-CoV-2.

**Methods:** This is an observational study generated from the first 79 patients admitted to Uppsala intensive care due to respiratory failure with SARS-CoV-2 infection. 34 underwent echocardiography of which 25 were included in the study and compared to 44 non-echo patients. Exclusion was based on absence of normofrequent sinus rhythm and mechanical respiratory support. Biomarker analysis was carried out on all patients.

**Results:** Mortality was increased in the echo compared to non-echo group (44 % vs. 16%, p<0.05). Right sided dimensions and functional parameters were not affected. Tricuspid regurgitation velocity indicated how increased pulmonary artery pressure was associated with mortality (survivors (n=5): 2.51 ± 0.01 m/s vs. non-survivors (n=5): 3.06 ± 0.11 m/s, p<0.05). Cardiac markers and D-dimer correlated to initiation of echocardiography (hs-TnI (ng/L): echo (n=23): 133 ± 45 vs. non-echo (n=41): 81.3 ± 45, p<0.01; NTproBNP (ng/L): echo (n=25): 2959 ± 573 vs. non-echo (n=42): 1641 ± 420, p<0.001; D-dimer (mg/L): echo (n=25): 16.1 ± 3.7 vs. non-echo (n=43: 6.1 ± 1.5, p<0.01) and mortality (hs-TnI (ng/L): survivors (n=48): 59.1 ± 21 vs. non-survivors (n=17): 211 ± 105, p<0.0001; NT-proBNP (ng/L): survivors (n=47): 1310 ± 314 vs. non-survivors (n=20): 4065 ± 740, p<0.0001; D-dimer (mg/L): survivors (n=50): 7.2 ± 1.5 vs. non-survivors (n=18): 17.1 ± 4.8, p<0.01). All intervals refer to standard error of the mean. Tricuspid regurgitation velocity was correlated with troponin I (r=0.93, r^2^=0.74, p<0.001, n=10).

**Conclusions:** These results suggest that there is no clear negative effect on cardiac function in critical SARS-CoV-2. There are indications that pulmonary pressure elevation carries a negative predictive outcome suggesting pulmonary disease as the driver of mortality. Cardiac biomarkers as well as D-dimer carry predictive value.

**Trial registration number:** Patients were included in “Clinical trials NCT04316884”

**Article summary:** *Strength and limitations of this study:*

- The patient body is recruited from all patients admitted to ICU in need of mechanical respiratory support independent of background which makes it relevant to clinical practice.
- The echocardiographic image acquisition was carried out by hospital assigned agents on clinical indication, which makes the results applicable in a clinical setting.
- Since the image acquisition was carried out on a clinical indication, the results may be skewed towards the false positive if applied to all Covid19 patients.

## Introduction

Since the severe acute respiratory syndrome corona virus 2 (SARS-CoV-2) was established as an infection transmitted amongst humans, it has been under intense scrutiny. The effects on the cardiovascular system are still not completely understood.

The respiratory tract is undisputed as the main target of SARS-CoV-2, but reports of myocarditis have been published (1), with autopsy findings showing myocardial T-cell infiltration (2, 3) and SARS-CoV-2 genome in myocardial biopsies from 104 patients with suspected myocarditis (4). Cellular internalisation through the angiotensin converting enzyme 2 (ACE-2) receptor also generated initial concern that heart failure and hypertension patients treated with angiotensin (Ang) inhibitors would be at increased risk due to upregulation of ACE-2 (5) and that increased ACE-2 shedding lowers the AngI/AngII causing vasoconstriction, inflammation, and risk of thrombosis (6). The occurrence of venous thromboembolic disease, as well as troponin and natriuretic peptides have been shown to correlate with negative outcomes (6). Thus, patient assessment by echocardiography has attracted clinical interest.

In a study of 305 patients, 62 % had myocardial injury as defined by elevated cardiac troponin which was associated with a both left (LV) and right (RV) ventricular abnormalities, higher admission rate to the intensive care unit (ICU), and mortality (7). In 74 patients with elevated troponin, 82 % requiring mechanical ventilation, mainly RV affliction associated with elevated D-dimer but not cardiac troponins, was observed. In the same cohort LV function was described as normal to hyperdynamic (8). In another study, LV dysfunction was not associated with higher mortality nor troponin levels, and RV dysfunction only present in 3/38 patients (9). In 18 patients stratified into mild and severe coronavirus disease 2019 (Covid-19) only the severely ill showed elevated measures and increased end-diastolic LV pressures without an effect on LV ejection fraction (LVEF) or RV function (10).

RV function has gained particular attention for the management of this disease. Right but not left sided affliction was associated with death in a cohort where 63 % of 94 patients were on mechanical ventilation support (11). In 120 patients RV function and pulmonary pressure, but not left sided parameters were identified as predictors of increased mortality (12). In 200 non-ICU-patients pulmonary hypertension (PH) without RV affliction was associated with a worsened outcome such as death or ICU admission (13).

We report from an exploratory study of Covid-19 patients admitted to the ICU of Uppsala University Hospital, the interrelationship between clinically initiated echocardiography, echocardiographic findings, biomarker levels, and mortality.

## Method

This study is a sub study of a prospective cohort study of patients that were admitted to the ICU at Uppsala University Hospital because of Covid-19. All patients were diagnosed by polymerase chain reaction from respiratory tract swabs and had respiratory failure requiring at least high flow oxygen therapy before admission to the ICU. Patients that were investigated by echocardiography were labelled “echo” patients and those that were not investigated by echocardiography were labelled “non-echo” patients.

The study was approved by the Swedish Ethical Review Authority (EPM; 2020-01623). The protocol of the study was registered (Clinical trials ID: NCT04316884). The declaration of Helsinki and its subsequent revisions were followed. Informed consent was given på subject patients.

Echocardiographic examination was carried out on clinically deemed indication by hospital certified sonographers. Analysis was carried out offline, independent of clinical analysis results, on TomTec^®^ software by the primary analyst and quality-controlled by a senior echocardiographer. Analyses of poor image quality were discarded at primary analyst’s discretion. Only patients with normo-frequent sinus rhythm were included for echocardiographic analysis. Pericardial effusion was quantified in the extra RV-RA space from the subcostal longitudinal view. For patients serially investigated by echocardiography, only the first examination was included. Normal values were assessed in relation to European Association of Cardiovascular Imaging (EACVI) consensus papers by Galderisi et al (14) and Lang et al (15). For assessment of pulmonary pressure, the 2015 European Society of Cardiology (ESC) guidelines were used (16).

Concentrations of clinically initiated biochemical cardiac blood markers during the stay in ICU, highly sensitive troponin I (hs-TnI), N-terminal pro brain natriuretic peptide (NT-proBNP), and D-dimer were compared between echo and non-echo investigated patients.

Biomarker values were correlated to separate echocardiographic parameters to investigate the predictive value of such parameters on cardiac function and strain. Concentrations in the survivor and non-survivor subgroups were compared to assess a possible predictive importance from increasing levels in serum.

Frequency distribution testing between groups were carried out using Chi-squared test or Fischer’s exact test for group sizes smaller than n=5. Groups were compared using Student’s t-test for normally distributed data and Mann-Whitney’s U-test for non-normally distributed data. Correlation analysis was performed using Pearson test for normally distributed data and Spearman’s test for non-normally distributed data. Bonferroni multiple analysis correction was applied to reduce the risk of type 1 errors, but pre-correction values are also reported. All confidence intervals are presented as standard error of the mean (SEM) unless otherwise stated. All statistical analyses and graphs utilised GraphPad Prism 5.0.

### Patient and public involvement

The study was carried out on patients in intensive care and a set of parameters investigated were established independently of patients’ priorities, experience and preferences. The design of the study was such that patients were involved as study subjects and not in the design of the study. Patients accepted involvement in the study through informed consent and served as study objects. They were not actively involved in recruitment and study conduct. Study results are available to patients on request and through publication.

## Results

### Patient selection

Out of 79 patients originally included in the study, 34 had been assessed by echocardiography as deemed indicated by the treating physician. Out of these 9 were subsequently excluded due to initial incorrect Covid-19 diagnosis, irregular heart rhythm, 3^rd^ degree AV-block, or overall unacceptable image quality, leaving 25 for analysis. 45 patients were included in the non-echo arm, of which one was excluded due to lack of respiratory insufficiency and oxygen therapy, leaving 44 patients in the non-echo group. A total of 69 patients were thus included in the study (fig 1).

**Figure 1:**
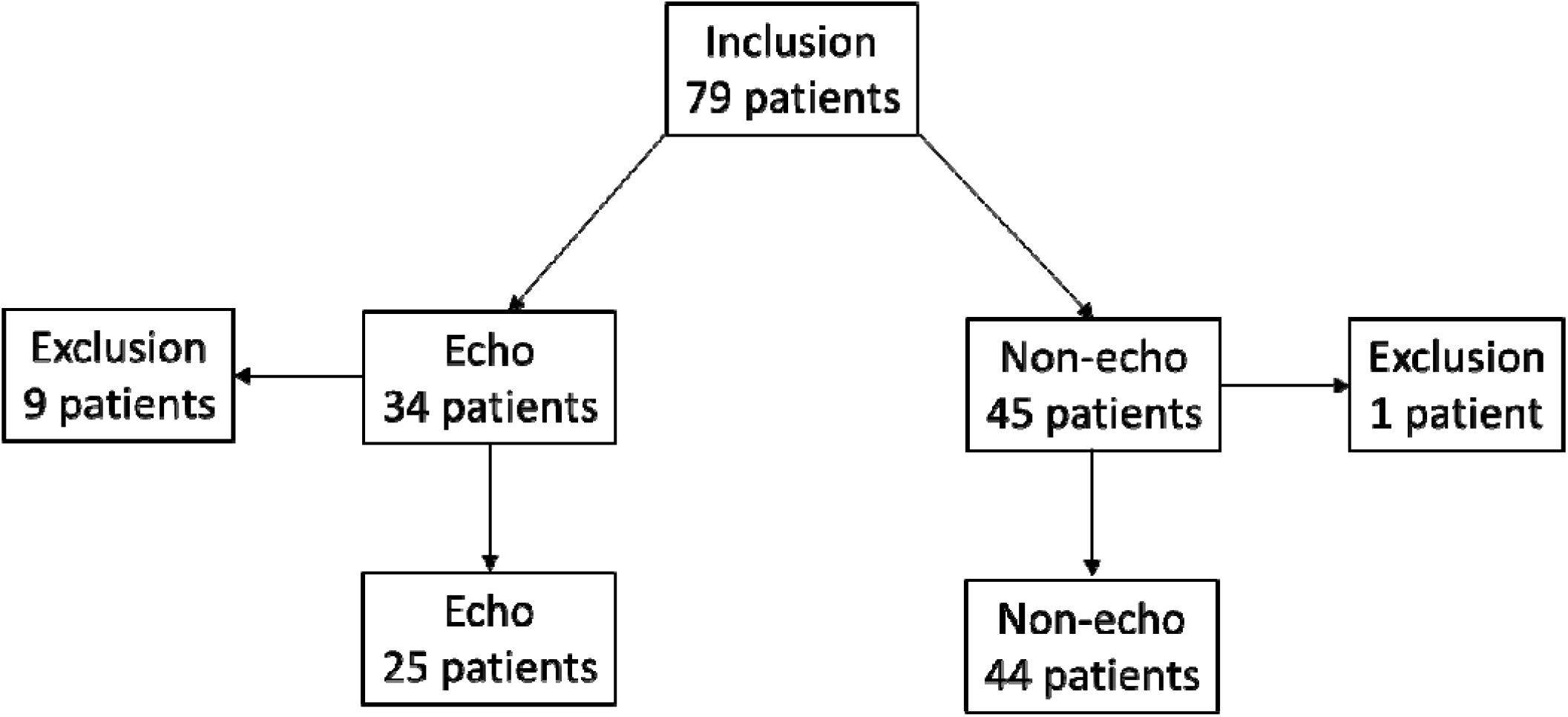
Out of 79 patients included in the study, 34 were assessed by echocardiography. Out of these 9 patients were subsequently excluded (3 did not fulfil the inclusion criteria of Covid-19 infection, 4 due to irregular heart rhythm or 3^rd^ degree AV-block, and 2 due to overall unacceptable image quality), leaving 25 valid for echocardiographic analyses and inclusion. 45 patients were not assessed by echocardiography, of which one was excluded (admitted to the intensive care unit due to concomitant diabetic ketoacidosis and not Covid-19 respiratory insufficiency), leaving 44 patients not assessed by echocardiography and a total of 69 patients in the study.

### Demographic characterisation

There were more male than female patients overall (54:15), this ratio was less pronounced in the echo group (19:6) vs the non-echo group (35:9). Mean age in the echo group (64.4 ± 2.6 years) was higher than in the non-echo group (56.7 ± 2.2 years) (p<0.05). BMI in the overall cohort was increased (28.9 ± 0.69 kg/m^2^) but there was no difference between the echo (28.8 ± 1.1 kg/m^2^) and non-echo group (29.0 ± 0.9 kg/m^2^) (p>0.05). Overall mortality in the study was 26 %, this was higher in the echo group (44 %) compared to the non-echo group (16 %) (p<0.05). Overall percentage of invasive ventilation was 55 %, also higher in the echo (80 %) group compared to in the non-echo group (41 %) (p<0.01). There was a higher presence of previous morbidities in the echo group (24%) than in the non-echo group (4.5 %) regarding ischemic heart disease (p<0.05) and preceding treatment with angiotensin converting enzyme inhibitors (ACEi) or angiotensin receptor blockers (ARB) which was higher in the echo group (52 %) compared to non-echo (25 %) (p<0.05). There was no difference in regards to previous heart failure, hypertension, pulmonary disease, diabetes mellitus type 2, or renal failure (table 1).

**Table 1:** Demographic characterisation. Morbidities refer to diagnoses previous to ICU admission. Confidence intervals described as standard error of the mean (SEM). Statistical analysis refers to non-echo vs echo group.

| **Parameter** | **Whole cohort** | **Non-echo** | **Echo** | **p-value** |
| --- | --- | --- | --- | --- |
| Gender (male:female) | 54:15 | 35:9 | 19:6 | ns |
| Age (years) | 59.5 ± 1.7 (n=69) | 56.7 ± 2.2 (n=44) | 64.4 ± 2.6 (n=25) | <0.05 |
| BMI (kg/m<sup>2</sup>) | 28.9 ± 0.69 (n=58) | 29.0 ± 0.9 (n=35) | 28.8 ± 1.1 (n=23) | ns |
| Mortality | 18/69 (26 %) | 7/44 (16 %) | 11/25 (44 %) | p<0.05 |
| Invasive ventilation | 38/69 (55 %) | 18/44 (41 %) | 20/25 (80 %) | p<0.01 |
| Ischemic heart disease | 8/69 (12 %) | 2/44 (4.5 %) | 6/25 (24%) | p<0.05 |
| Heart failure | 3/69 (4.3 %) | 1/44 (2.3 %) | 2/25 (8 %) | ns |
| Pulmonary disease | 14/69 (20 %) | 7/44 (16 %) | 7/25 (28 %) | ns |
| Hypertension | 34/69 (49 %) | 19/44 (43 %) | 15/25 (60 %) | ns |
| Diabetes mellitus | 19/69 (28 %) | 10/44 (23 %) | 9/25 (36 %) | ns |
| Renal failure | 18/69 (26 %) | 15/44 (34 %) | 3/25 (12 %) | ns |
| ACEi/ARB | 24/69 (35 %) | 11/44 (25 %) | 13/25 (52 %) | p<0.05 |

### Biomarkers

Admission level of hs-TnI was higher in the non-survivor group (p<0.01) and D-dimer slightly but significantly lower in the non-survivor group (p<0.05) (table 2 a). Maximum expression levels of hs-TnI (p<0.0001), NT-proBNP (p<0.0001) and D-dimer (p<0.01) were all significantly higher in the non-survivor group (table 2 b). Maximum expression levels of hs-TnI (p<0.01), NT-proBNP (p<0.001), and D-dimer (p<0.01) was higher in the echo group compared to non-echo group (table 2 c).

**Table 2 a:** Admission biomarker level’s relation to mortality. Confidence intervals described as standard error of the mean (SEM). Abbreviations: Highly sensitivity troponin I (hs-TnI), N-terminal brain natriuretic peptide (NT-proBNP), not significant (ns).

| **Marker** | **Whole cohort** | **Survivors** | **Non-survivors** | **p-value** |
| --- | --- | --- | --- | --- |
| hs-TnI (ng/L) | 85.4 ± 41 (n=47) | 39.4 ± 15 (n=32) | 180 ± 120 (n=15) | <0.01 |
| NT-proBNP (ng/L) | 1055 ± 276 (n=48) | 936 ± 351 (n=33) | 1318 ± 438 (n=15) | ns |
| D-dimer (mg/L) | 2.58 ± 0.6 (n=63) | 2.64 ± 0.6 (n=46) | 2.42 ± 0.4 (n=17) | <0.05 |

**Table 2 b:** Maximum biomarker level’s relation to mortality. Confidence intervals described as standard error of the mean (SEM).

| **Marker** | **Whole cohort** | **Survivors** | **Non-survivors** | **p-value** |
| --- | --- | --- | --- | --- |
| hs-TnI (ng/L) | 98.8 ± 32 (n=65) | 59.1 ± 21 (n=48) | 211 ± 105 (n=17) | <0.0001 |
| NT-proBNP (ng/L) | 2133 ± 346 (n=67) | 1310 ± 314 (n=47) | 4065 ± 740 (n=20) | <0.0001 |
| D-dimer (mg/L) | 9.8 ± 1.8 (n=68) | 7.2 ± 1.5 (n=50) | 17.1 ± 4.8 (n=18) | <0.01 |

**Table 2 c:** Biomarker distribution in the non-echo vs. echo group. Confidence intervals described as standard error of the mean (SEM).

| **Marker** | **Non-echo** | **Echo** | **p-value** |
| --- | --- | --- | --- |
| hs-TnI (ng/L) | 81.3 ± 45 (n=41) | 133 ± 45 (n=23) | <0.01 |
| NT-proBNP (ng/L) | 1641 ± 420 (n=42) | 2959 ± 573 (n=25) | <0.001 |
| D-dimer (mg/L) | 6.1 ± 1.5 (n=43) | 16.1 ± 3.7 (n=25) | <0.01 |

Correlation between standard echocardiographic parameters and maximum biomarker levels was investigated showing that hs-TnI was positively correlated to maximum tricuspid valve regurgitation velocity (TR V_max_) (p<0.01) (fig 2). There were no significant correlations between other cardiac biomarkers or D-dimer and specific echocardiographic parameters after multiple comparison correction that stood up to multiple comparison correction (supplementary data table 1).

**Figure 2:**
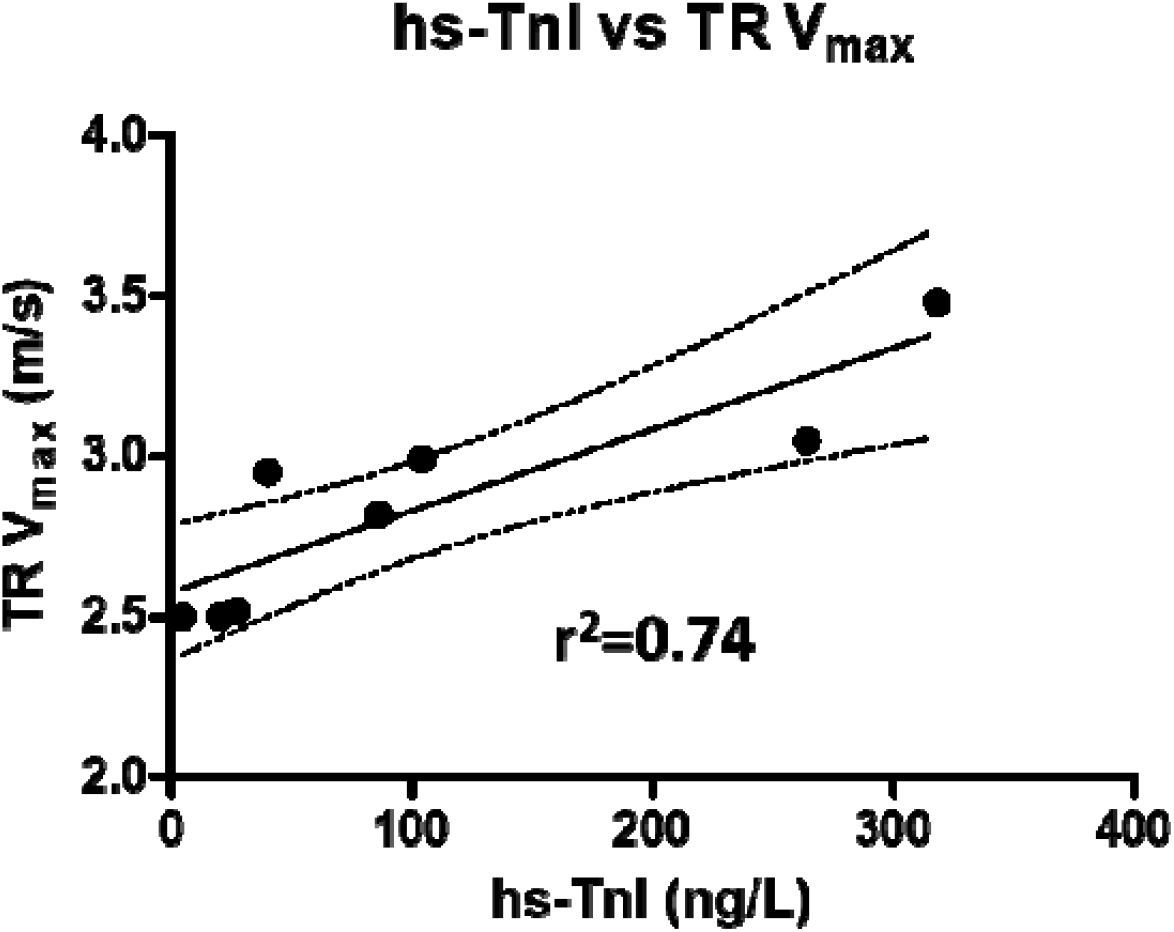
Significant predictive value of maximum high sensitivity troponin I (hs-TnI) in relation to maximum tricuspid valve regurgitation velocity (TR V_max_). Linear regression fit with 95% confidence intervals and best fit. For p- and r-values, see supplementary data table 1.

### Echocardiography

In the overall echocardiographic assessment, all four heart chambers were of normal size as of average values, LA/m^2^ (23.8 ± 1.61 ml/m^2^ (<34), of which 3/20 exceeded normal volume/m^2^), LVEDD (47.7 ± 1.3 mm (<58.4 (male), <52.2 (female) of which 2/22 exceeded normal diameter), RA/m^2^ (24.2 ± 2.9 ml/m^2^ (<30 (male), <28 (female) of which 7/18 exceeded normal volume/m^2^), apart from RVD1 (36.1 ± 1.5 mm (<36), of which 11/22 exceeded normal values (14)). The interventricular septal diameter (IVSD) average was slightly increased (11.4 ± 0.5 mm (6-10 (male) 6-9 (female), of which 15/22 exceeded normal values) as well as posterior wall thickness (10.1 ± 0.3 mm (6-10 (male), 6-9 (female), of which 8/22 exceeded normal values (15). Average LV global longitudinal strain (GLS) was reduced (−14.1 ± 0.86 % (<-20), low in 17/18 patients, despite a normal LVEF (54.8 ± 2.2 % (>52 (male), >54 (female), which was low in 5/20 patients, of which 3 had a previous diagnosis of heart failure or coronary disease). Average RV systolic function, judging by TAPSE (22.7 ± 1.1 mm (>17), reduced in 3/22), RV fractional area change (RV FAC) (45.2 ± 2.4 % (>35), reduced in 4/20 patient) and RV free wall strain (−25.1 ± 2.5 % (<-23), below normal in 8/16 patients) was within normal range (14). Transmitral early diastolic velocity was increased (0.77 ± 0.04 m/s (<0.5), exceeding normal in 17/20 patients), but E/A ratio (1.30 ± 0.13 (0.8-2.0), abnormal in 4/20 patients), septal e’ (0.079 ± 0.01 m/s (>0.07), reduced in 8/16 patients), E/e’ (10.5 ± 0.81 (<14), increased in 4/16 patients) as well as transmitral deceleration time (MV dec. time) (211 ± 12 ms (160-220), abnormal in 11/20 patients) were normal (14). IVC dimension (20.0 ± 0.84 mm (<20), increased in 10/18 patients) and its respiratory variation (49.5 ± 7.3 % (>50), reduced in 10/17) failed to indicate clear signs of increased RA pressure (14). Average pulmonary artery acceleration time (PAAT) (109 ± 6.3 ms (<100 ms, highly suggestive of PH (6/16 patients), 100-130 ms, intermediate probability of PH (7/16 patients)) (17), and maximum tricuspid regurgitation velocity (TR V_max_) (2.78 ± 0.11 m/s (<2.8 m/s), elevated in 5/10 patients, were normal (14). Non-survivors in ICU had a pre multiple comparison-correction higher TR V_max_ (3.06 ± 0.11 ms) than survivors (2.51 ± 0.01 m/s) (14), however losing significance after correction. There were no increased amounts of pericardial effusion (0.71 ± 0.25 mm) (mild<10 mm (no patients with more than 10 mm) (18). Apart from TR V_max_ there were no other parameters that differed between the survivor and non-survivor group (table 3). None of the scanned patients had any marked valvular stenoses or regurgitations.

**Table 3:** Echocardiographic parameters in the whole group and their distribution between survivor and non-survivor group. Confidence intervals described as standard error of the mean (SEM). Statistical analyses refer to survivor vs. non-survivor group.

| **Parameter** | **Whole group** | **Survivors** | **Non-survivors** | **p-value (Bonferroni correction)** |
| --- | --- | --- | --- | --- |
| <u>Static measures</u> |  |  |  |  |
| IVSD (mm) | 11.4 ± 0.5 (n=22) | 11.7 ± 0.58 (n=12) | 10.9 ± 0.88 (n=10) | ns |
| LVEDD (mm) | 47.7 ± 1.3 (n=22) | 45.9 ± 1.8 (n=12) | 49.8 ± 1.6 (n=10) | ns |
| LVEDD/m<sup>2</sup> (mm/mm<sup>2</sup>) | 23.78 ± 0.57 (n=22) | 23.3 ± 0.90 (n=12) | 24.4 ± 0.62 (n=10) | ns |
| LVPWD (mm) | 10.1 ± 0.3 (n=22) | 10.2 ± 0.4 (n=12) | 9.8 ± 0. (n=10) | ns |
| LA (ml) | 47.8 ± 3.3 (n=20) | 44.1 ± 3.7 (n=11) | 52.4 ± 5.5 (n=9) | ns |
| LA/m<sup>2</sup> (ml/m<sup>2</sup>) | 23.8 ± 1.61 (n=20) | 22.3 ± 1.6 (n=11) | 25.7 ± 3.0 (n=9) | ns |
| RVD1 (mm) | 36.1 ± 1.5 (n=22) | 34.2 ± 2.0 (n=12) | 38.4 ± 2.0 (n=10) | ns |
| RVD1/m<sup>2</sup> (mm/m<sup>2</sup>) | 18.0 ± 0.61 (n=22) | 17.43 ± 0.85 (n=12) | 18.7 ± 0.86 (n=10) | ns |
| RA (ml) | 49.0 ± 6.1 (n=18) | 46.4 ± 8.2 (n=10) | 52.1 ± 9.6 (n=8) | ns |
| RA/m<sup>2</sup> (ml/m<sup>2</sup>) | 24.2 ± 2.9 (n=18) | 23.7 ± 3.9 (n=10) | 24.9 ± 4.5 (n=8) | ns |
| IVC diameter (mm) | 20.0 ± 0.84 (n=18) | 19.0 ± 1.4 (n=7) | 20.6 ± 1.1 (n=11) | ns |
| Pericardial effusion (mm) | 0.71 ± 0.25 (n=24) | 0.97 ± 0.39 (n=13) | 0.41 ± 0.29 (n=11) | ns |
| <u>Functional measures</u> |  |  |  |  |
| LVEF (%) | 54.8 ± 2.2 (n=20) | 54.1 ± 3.1 (n=11) | 55.7 ± 3.4 (n=9) | ns |
| GLS (%) | -14.1 ± 0.86 (n=18) | -13.5 ± 1.3 (n=11) | -15.1 ± 1.0 (n=7) | ns |
| E (m/s) | 0.77 ± 0.04 (n=20) | 0.71 ± 0.05 (n=10) | 0.83 ± 0.07 (n=10) | ns |
| A (m/s) | 0.64 ± 0.05 (n=20) | 0.62 ± 0.05 (n=10) | 0.67 ± 0.05 (n=10) | ns |
| E/A | 1.30 ± 0.13 (n=20) | 1.27 ± 0.23 (n=10) | 1.33 ± 0.13 (n=10) | ns |
| e' medial (m/s) | 0.079 ± 0.01 (n=16) | 0.072 ± 0.01 (n=6) | 0.083 ± 0.01 (n=10) | ns |
| E/e' | 10.5 ± 0.81 (n=16) | 9.72 ± 0.54 (n=6) | 10.9 ± 1.3 (n=10) | ns |
| MV dec time (ms) | 211 ± 12 (n=20) | 203 ± 21 (n=10) | 220 ± 11 (n=10) | ns |
| TAPSE (mm) | 22.7 ± 1.1 (n=22) | 23.4 ± 1.5 (n=12) | 21.8 ± 1.6 (n=10) | ns |
| RV FAC (%) | 45.2 ± 2.4 (n=20) | 47.9 ± 2.4 (n=10) | 42.4 ± 4.1 (n=10) | ns |
| RV free wall strain (%) | -25.1 ± 2.5 (n=16) | -26.3 ± 3.5 (n=9) | -23.6 ± 3.6 (n=7) | ns |
| IVC resp. variation (%) | 49.5 ± 7.3 (n=17) | 59.4 ± 11.2 (n=7) | 42.6 ± 9.5 (n=10) | ns |
| PAAT (ms) | 109 ± 6.3 (n=16) | 115 ± 9.2 (n=9) | 101 ± 8.0 (n=7) | ns |
| TR V<sub>max</sub> (m/s) | 2.78 ± 0.11 (n=10) | 2.51 ± 0.01 (n=5) | 3.06 ± 0.11 (n=5) | <0.05 (0.3) |

## Discussion

The non-invasive nature and high availability of transthoracic echocardiography makes it an attractive method of assessing critically ill patients in intensive care that are often in need of serial investigations to interpret effects of treatments on the underlying condition. During spring 2020 an increasing number of patients were admitted to intensive care due to Covid-19 respiratory failure. The lungs are the main target organ of this infection, but there have been reports of myocarditis and effects on both LV and RV function which is theoretically plausible through cellular internalisation via myocardial ACE-2 receptors and indeed, SARS-CoV-2 myocarditis including myocardial virus replication has now been reported (4).

From patients admitted to the ICU at Uppsala University Hospital during the first wave of Covid-19 we attempted to investigate which parameters that are affected in this group and also to derive a predictive value from the echocardiographic examination. We also correlated standard cardiac biomarkers and D-dimer values at admission as well as the maximum values in ICU to outcome and echocardiographic parameters.

The mortality in the group investigated by echocardiography was higher than those that were not assessed which suggests that these patients exhibited a physiological and biochemical status of increased cardiopulmonary strain which explains the higher mortality. For overall assessment of the whole cohort, the LV systolic function as assessed by GLS was lower in these patients than what to expect from standard population material (14). The image quality as well as patient cooperativity was however reduced in these patients compared to standard reference materials which confounds the interpretation of this isolated parameter as evidence of myocardial damage. The LVEF was not affected on average, and 3/5 patients with LVEF less than normal had a previous diagnosis of heart failure or coronary disease, neither were there signs of increased amounts of pericardial effusion. Overall diastolic dysfunction as assessed by combined evaluation of E/e’, isolated septal diastolic movement velocity (e’), LA volume, and TR V_max_ could not be verified (19). Thus, there were no overt echocardiographic signs of myocarditis in these patients.

Systolic and diastolic function of the LV, as judged by LVEF, GLS, and e’, showed no association to mortality, but there were insignificant trends towards increased LV end-diastolic diameter and LA size in the non-survival group. IVSD and left ventricle posterior wall end diastolic diameter (LVPWD) were both slightly thicker than what to expect from reference materials, this most likely reflects a high average age and presence of hypertension in the studied cohort, and trended towards thinning in the non-survival group. These findings could suggest fluid overload and passive dilation in this group, which is supported by the increased level of NT-proBNP, thus providing no evidence of myocarditis-induced oedema. Right sided parameters indicated a trend towards association with mortality. Systolic function as assessed by TAPSE, RV FAC, and RV free wall strain, showed an overall trend towards reduction in the non-survival group and the RVD1 was also increased to wider than 35 mm in 11/22 readings of all investigated patients, being insignificantly increased to 38.4 ± 2.0 mm in the non-survivor group compared to 34.2 ± 2.0 mm in the survivor group, which may indicate increased afterload of the RV. TR V_max_ was increased in the non-survivor group but failed to show significance after multiple analysis correction, which suggests an elevated pressure in the pulmonary circulation. Average PAAT was also found to be in the intermediate strata suggestive of PH. In the absence of signs of LV diastolic impairment and pulmonary embolism diagnosed in only 4/25 echo patients, it is suggested this is due to hypoxia in the pulmonary arterial bed, pulmonary vasoconstriction, and RV strain.

Unsurprisingly, maximum levels of hs-TnI, NT-proBNP and D-dimer were associated with death. More interestingly, only increased admission level of hs-TnI was associated with mortality, with a trend for NT-proBNP. hs-TnI was also strongly associated with TR V_max_, suggesting from biochemical and echocardiographic data this parameter to be of importance in determining the outcome of Covid-19 patients in the ICU.

The main weakness of the study in detecting cardiac affliction from SARS-CoV-2, is that not all ICU patients underwent echocardiography, but only those where the treating physician found it indicated. This leads to a selection bias but the circumstance also enabled the comparison of echo vs. non-echo patients. However, this selection ought to produce a cohort with more severe cardiac dysfunction, meaning that our results may be over-estimating the degree of cardiac dysfunction rather than underestimating it. In addition, the small cohort size and large set of variables increases the risk of false positive effects, again suggesting that the study should tend to over-estimate the effect of critical Covid-19 on cardiac function, which was corrected for in the analysis.

The situation of echocardiography in the ICU-setting of reduced patient cooperativity and ventilation with positive intrathoracic pressure, will reduce availability of parameters, image quality and sensitivity of interpretation which risks failing to register significant associations. For GLS measurements 4/18 were made from 2/3 standard projections, for LVEF 2/20 were made from 1/2 standard projections and for LA volume 8/20 were made from 1/2 standard projections, since images of sufficient quality was not available.

In conclusion, there are no convincing signs of cardiac function being systematically affected in Covid-19 patients admitted to ICU, and no evidence that cardiac dysfunction is a major driver of mortality in critically ill patients with Covid-19. Still, cardiac biomarkers and D-dimer carry a predictive value in outcome, but more likely reflect strain on the heart from a primary pulmonary affliction.

## Key messages

### What is already known about this subject?

Covid-19 is primarily an infection that affects the lower respiratory tract but cases have been found of SARS-CoV-2 affecting the myocardium inducing myocarditis.

### What does this study add?

The presented study offers a review of intensive care admitted-patients requiring mechanical ventilation; findings from echocardiographic examination, the prognostic impact of clinically initiated echocardiography, and relevance of cardiac biomarkers and d-dimer. The material is, apart from this, unselected and therefore reflects what clinicians are likely to encounter giving care to this group of patients. It shows how from this group of patients, primary myocardial affliction is not a standard or common finding.

### How might this impact on clinical practice?

The need for echocardiography is a negative prognostic factor. An increased pulmonary pressure as assessed by maximum tricuspid valve regurgitation velocity suggests a negative outcome. Troponin I is significantly correlated to maximum tricuspid valve regurgitation velocity.

## Data Availability

Data is available from the author upon reasonable request

## Acknowledgements

The authors thank Research nurses Joanna Wessbergh and Elin Söderman, and biobank research assistants Labolina Spång, Erik Danielsson and Philip Karlsson for their expertise in compiling the study. We are also thankful to patient advisers with regards to recruitment and answering to questions regarding participation.

## Authorship contributions

HI: Carried out the primary echo image and data analysis and drafted the manuscript

AL: Provided facilities for biomarker analysis and reviewed the manuscript draft

ML: Recruited patients, and reviewed the manuscript draft

RF: Recruited patients, and reviewed the manuscript draft

FF: Controlled the echo analysis and reviewed the manuscript draft

MH: Conceptualised and designed the study, recruited patients and reviewed the manuscript draft

## Data availability

Data is available from the corresponding author on reasonable request.

## Supplementary data

**Table 1:** Correlation of echo parameters to cardiac biomarkers and d-dimer. (p-value, r, Bonferroni-corrected p-value). Abbreviations: Interventricular septum diameter (IVSD), left ventricular end diastolic diameter (LVEDD), left ventricular posterior wall end diastolic diameter (LVPWD), left atrium (LA), right ventricular basal diameter (RVD1), right atrium (RA), inferior vena cava (IVC), left ventricular ejection fraction (LVEF), global longitudinal strain (GLS), early transmitral velocity (E), atrial transmitral velocity (A), medial tissue diastolic velocity (e’), mitral valve deceleration time (MV dec. time), tricuspid annular peak systolic excursion (TAPSE), right ventricular fractional area change (RV FAC), right ventricular free wall strain (RV free wall strain), inferior vena cava respiratory variation (IVC resp. variation), pulmonary artery acceleration time (Pulm. Acc. Time), maximum tricuspid regurgitation velocity (TR V_max_), not significant (ns)

| **Parameter** | **hs-TnI (n)** | **NT-proBNP (n)** | **d-dimer (n)** |
| --- | --- | --- | --- |
| IVSD (mm) | <0.05, 0.46, 1.04 (n=20) | ns | ns |
| LVEDD (mm) | ns | ns | ns |
| LVEDD/m<sup>2</sup> (mm/m<sup>2</sup>) | ns | ns | ns |
| LVPWD (mm) | ns | ns | ns |
| LA (ml) | ns | ns | ns |
| LA/m<sup>2</sup> (ml/m<sup>2</sup>) | ns | ns | ns |
| RVD1 (mm) | ns | ns | ns |
| RVD1/m<sup>2</sup> (mm/m<sup>2</sup>) | ns | ns | ns |
| RA (ml) | ns | ns | ns |
| RA/m<sup>2</sup> (ml/mm<sup>2</sup>) | ns | ns | ns |
| IVC diameter (mm) | ns | ns | ns |
| LVEF (%) | ns | ns | ns |
| GLS (%) | ns | ns | <0.01, -0.62, 0.165 (n=18) |
| E (m/s) | ns | <0.05, 0.50, 0.59 (n=20) | <0.05, 0.45, 1.11 (n=20) |
| A (m/s) | ns | ns | ns |
| E/A | ns | ns | ns |
| e' medial (m/s) | ns | ns | ns |
| E/e' medial | ns | ns | ns |
| MV dec. time (ms) | ns | ns | ns |
| TAPSE (mm) | ns | ns | ns |
| RV FAC (%) | ns | <0.05, -0.45, 1.1 (n=20) | ns |
| RV free wall strain | ns | ns | ns |
| IVC resp. variation (%) | ns | ns | ns |
| Pulm. Acc. Time (ms) | ns | ns | ns |
| TR V<sub>max</sub> (m/s) | <0.001, 0.93, 0.0075 (n=10) | ns | ns |
| Pericardial effusion (mm) | ns | ns | ns |

## List of abbreviations

A: atrial transmitral diastolic velocity
ACE-2: angiotensin converting enzyme 2
ACEi: angiotensin converting enzyme inhibitor
ARB: angiotensin receptor blocker
Ang: angiotensin
BMI: body mass index
Covid-19: corona virus disease 2019
E: early transmitral diastolic velocity
e’: medial tissue diastolic velocity
EACVI: European association of cardiovascular imaging
ESC: European society of cardiology
FAC: fractional area change
GLS: global longitudinal strain
hs-TnI: high sensitivity troponin I
ICU: intensive care unit
IVC: inferior vena cava
IVC resp. variation: inferior vena cava respiratory variation
IVSD: interventricular septum diameter
LA: left atrium
LV: left ventricle
LVEDD: left ventricular end diastolic diameter
LVEF: left ventricular ejection fraction
LVPWD: left ventricular posterior wall end diastolic diameter
MV dec. time: mitral valve deceleration time
ns: not significant
NT-proBNP: N-terminal pro brain natriuretic peptide
PH: pulmonary hypertension
PAAT: pulmonary artery acceleration time
RA: right atrium
RV: right ventricle
RVD1: right ventricular basal diameter
SARS-CoV-2: severe acute respiratory syndrome corona virus 2
TAPSE: tricuspid annular peak systolic excursion
TR V_max_: maximum tricuspid regurgitation velocity

